# The role of physical activity in shaping the relationship between alcohol use and anxiety and depression symptoms: Findings from a U.S. nationwide cohort

**DOI:** 10.64898/2026.07.30.26359355

**Authors:** Jenna Sanborn, Denis Nash, McKaylee Robertson, Angela Parcesepe, Zachary Shahn

## Abstract

**Background:** Alcohol use and physical activity are common, interrelated behaviors with opposing associations to mental health, yet little is known about their joint relationship with anxiety and depression. This study evaluated whether the association between alcohol use and anxiety and/or depression symptoms differed by physical activity level in a large U.S. longitudinal cohort.

**Methods:** Using data from 5,152 adults in the CHASING COVID Cohort (2021-2023), alcohol use was categorized as low-, moderate-, or high-risk based on repeated AUDIT-C assessments, and physical activity was classified as active versus sedentary using items adapted from the 2023 Behavioral Risk Factor Surveillance System. Moderate to severe symptoms of anxiety/depression, assessed subsequent to the exposure assessment period, was defined as a PHQ-8 or GAD-7 score ≥10. Log-binomial regression estimated adjusted risk ratios (aRRs) for associations between alcohol use and anxiety/depression symptoms within strata of physical activity. Effect modification by physical activity was evaluated on the additive and multiplicative scales. Analyses were repeated among healthier participants without chronic conditions or poor self-rated health to reduce sick-quitter bias (N=3,141).

**Results:** Most participants had low-risk or no alcohol use (70.3%), followed by moderate-risk (21.9%) and high-risk use (7.8%); 29.5% were sedentary. Sedentary participants had higher risk of moderate to severe anxiety/depression symptoms than active participants across alcohol use levels overall and in the healthier subsample. In the overall sample, active participants with high-risk alcohol use had elevated risk compared with active participants with low-risk or no alcohol use (aRR = 1.44, 95% CI: 1.15-1.80) and physical activity did not modify the association between alcohol use and anxiety/depression symptoms on either scale. Among healthier participants, risk was highest among sedentary participants with moderate-risk alcohol use compared with active participants with low-risk or no alcohol use (aRR = 1.92, 95% CI: 1.50-2.45); physical activity modified the association between moderate-risk alcohol use and moderate to severe anxiety/depression symptoms on the additive scale (RERI = 0.61, p=0.013) and multiplicative scales (ratio of aRRs = 1.49, p=0.001).

**Conclusions:** Among healthier adults, physical activity modified the association between moderate alcohol use and symptoms of anxiety or depression on additive and multiplicative scales. Findings suggest sedentary behavior may amplify mental health vulnerability associated with moderate drinking; future studies are warranted to clarify these relationships, including those with larger numbers of high-risk drinkers.

## Introduction

Anxiety and depression are among the most common and disabling mental health conditions in the United States (U.S.), affecting more than one in five adults each year.^1^ These disorders frequently co-occur, are associated with increased risk of chronic disease and premature mortality,^2–10^ and rank among the leading causes of global disability.^11,12^ Alcohol use and physical activity are two common behaviors shown to be associated with mental-health outcomes. Alcohol use has been positively associated with anxiety and depressive disorders, particularly among heavy drinkers.^13–18^ Systematic reviews support a bi-directional relationship, whereby anxiety and depression may contribute to alcohol use, while alcohol use may exacerbate symptoms through neurophysiological, biological and social pathways.^16–18^ In contrast, experimental and observational studies consistently show that physical activity is associated with lower symptoms of anxiety and depression across diverse populations and activity levels.^19–26^

Although alcohol use and physical activity have divergent associations with anxiety and depression, they often co-occur. Population-based studies show that adults with higher activity levels are more likely to drink at moderate and high levels than less active adults, even after adjustment for sociodemographic and health factors.^27–32^ In a population-representative study of U.S. adults, light, moderate, and heavy drinkers exercised an average of 6, 10 and 20 more minutes per week, respectively, than abstainers after adjustment for various factors.^31^ This clustering may contribute to inconsistent findings in alcohol-mental health research, where observational studies commonly report a J- or U-shaped association, with low-risk and moderate drinkers appearing to have better mental health outcomes than abstainers and heavy drinkers.^33–42^ These patterns may partly reflect bias from health-related selection into abstinence (sick-quitter bias),^43–46^ as well as inadequate adjustment for physical activity, which has often been omitted from studies of alcohol use and depressive symptoms.^40^

Together, existing literature suggests that alcohol use, physical activity, and mental health are behaviorally and causally intertwined, yet little is known about whether physical activity modifies the association between alcohol use and anxiety or depression symptoms. It has been hypothesized that physical activity may mitigate harmful mental health effects of alcohol use among drinkers, but this possibility has not been investigated.^27,31,47–49^ In this study, we used data from the CHASING COVID Cohort Study to assess whether physical activity modifies the association between alcohol use and symptoms of anxiety or depression, including whether alcohol use and sedentary behavior were jointly associated with excess symptom burden beyond their independent associations.^40^

We evaluated effect modification on both additive and multiplicative scales. Additive-scale effect modification assessed whether the joint association exceeded the sum of the individual associations, while multiplicative-scale effect modification assessed whether the relative association between alcohol use and anxiety/depression symptoms differed by physical activity level. We hypothesized that alcohol-related risk of anxiety and depression symptoms would be stronger among sedentary participants than among physically active participants, consistent with a potential protective role of physical activity among moderate to heavy drinkers. We further hypothesized that the joint association of alcohol use and sedentary behavior on anxiety and depression symptoms would exceed the sum of their individual associations, indicating greater overall mental health burden among individuals who both consume alcohol above low-risk levels and are physically inactive. We examined associations in the overall sample and among participants without chronic conditions or poor self-rated health to reduce the influence of sick-quitter bias. This descriptive analysis aims to generate evidence on the interrelationship between these behaviors and symptoms of depression and anxiety and inform future causal investigations and intervention strategies focused on modifiable behavioral risks.

## Methods

### Data source

The Communities, Households, and SARS-CoV-2 Epidemiology (CHASING) COVID Cohort study has followed 5,798 socio-demographically diverse adults (aged 18+) residing across all 50 U.S. states, Washington D.C., Puerto Rico, and Guam since 2020. The study collected longitudinal data on a broad range of health-related behaviors and outcomes. Participants were recruited online, and follow-up assessments were administered approximately quarterly through December 2023. Recruitment procedures have been previously described^50^ and study assessments are accessible online.^51^

This study was approved by the Institutional Review Board at the City University of New York Graduate School for Public Health and Health Policy, and participants could voluntarily discontinue participation at any time. Participant consent for participation was obtained at baseline and at periodic follow-up assessments.

### Analytic sample

This study focused on data collected after September 2021, when COVID-19 vaccines were widely available and approved for all U.S. adults aged 18 years and older. Restricting analyses to this post-vaccine period allowed evaluation of the mental health relationship with behavioral factors after the acute phase of the pandemic had stabilized, minimizing the influence of stressors specific to the early lockdown period. The analytic sample included participants who completed at least one survey with alcohol consumption data from September 2021-September 2023 (September 2021, March 2022, October 2022, April 2023, and September 2023; N = 5,152). Among these, 77% completed all five surveys, and 91% completed at least three.

### Alcohol use (exposure)

Alcohol use was measured using the 3-item Alcohol Use Disorders Identification Test-Consumption (AUDIT-C), a validated screening tool recommended by the U.S. Preventive Services Task Force for detecting unhealthy alcohol use. The AUDIT-C sums three items assessing drinking frequency, typical quantity, and frequency of heavy drinking, yielding scores ranging from 0 (no use) to 12.^52^ Consumption was categorized into three levels-low risk, moderate risk, and high-risk-based on sex-specific cutoff scores applied across 5 survey waves, following guidelines adapted from the U.S. Department of Veterans Affairs (Table 1).^53^ For males, scores of 0-3 indicated low risk, 4-5 moderate risk, and ≥6 high/severe risk; for females, scores of 0-2 indicated low risk, 3-5 moderate risk, and ≥6 high/severe risk. Participants were classified as persistently high-risk if ≥50% of assessments indicated high-risk use, persistently low-risk if ≥50% indicated low-risk use, and moderate-risk otherwise.

**Table 1.**
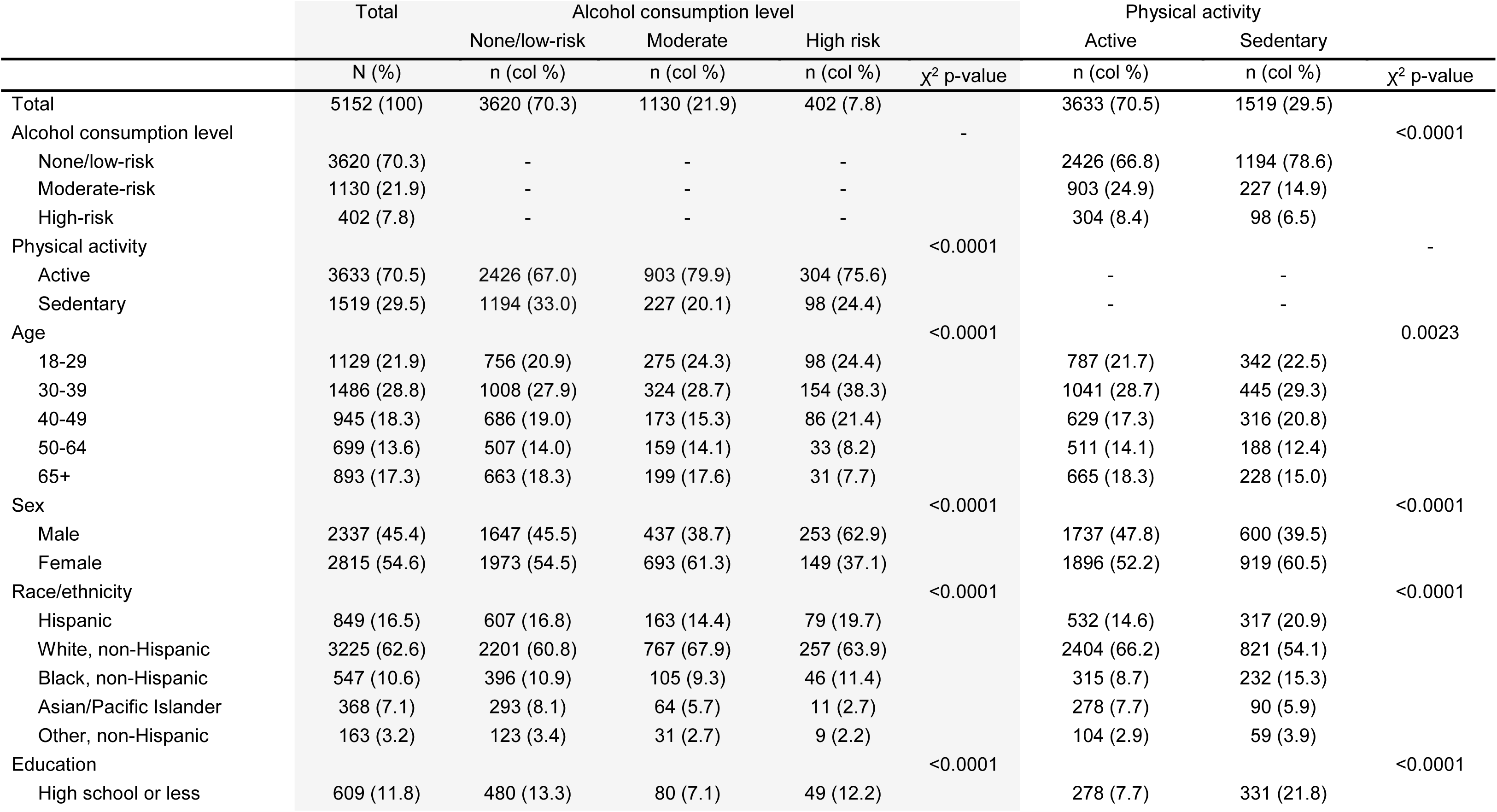
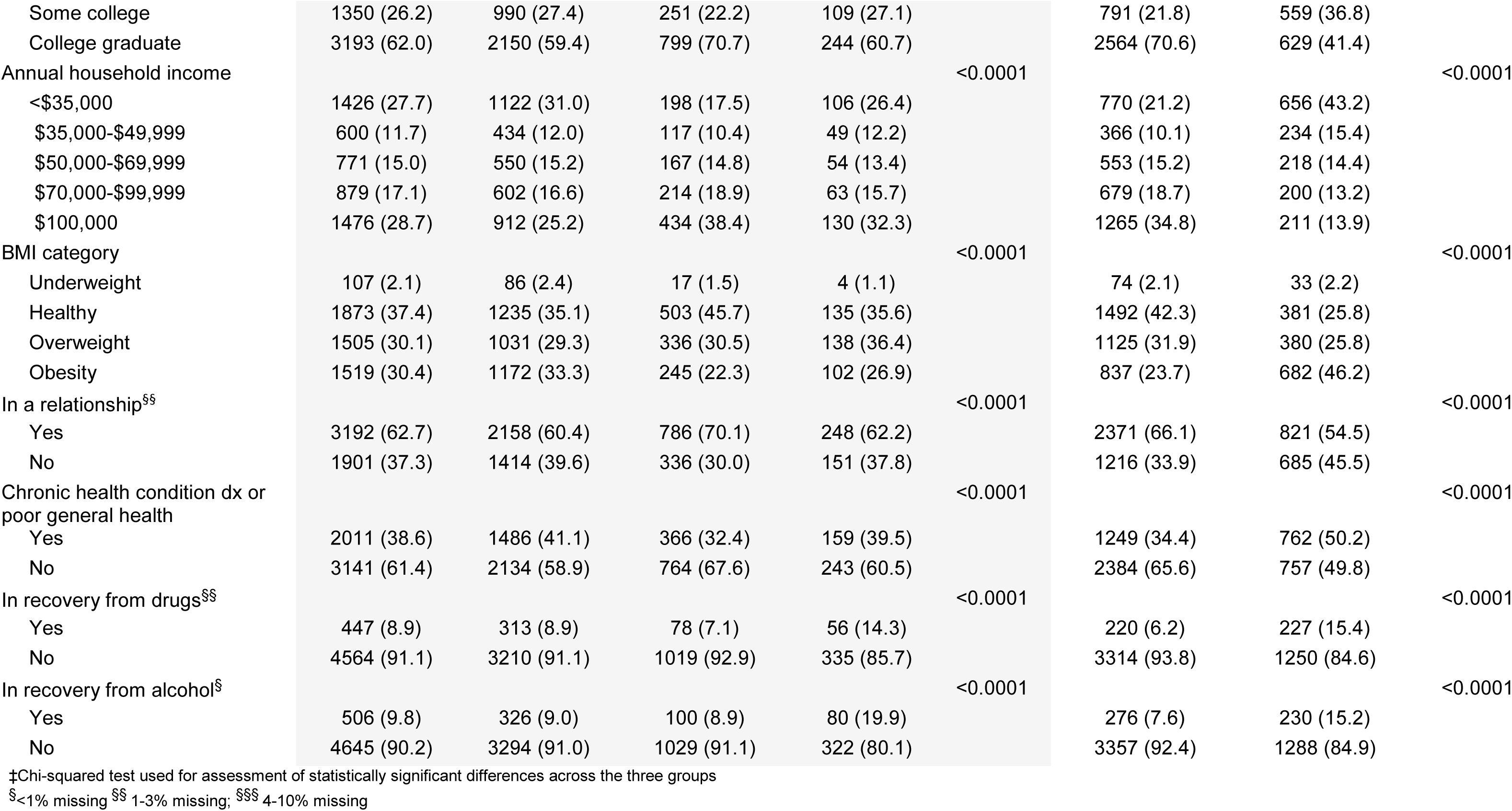
Cohort characteristics by exposure and modifier status, CHASING COVID Cohort, September 2021-December 2023 (N=5,152)

| | Total | Alcohol consumption level | | | $\chi^2$ p-value | Physical activity | | $\chi^2$ p-value |
| --- | --- | --- | --- | --- | --- | --- | --- | --- |
|  | N (%) | None/low-risk<br>n (col %) | Moderate<br>n (col %) | High risk<br>n (col %) |  | Active<br>n (col %) | Sedentary<br>n (col %) |  |
| Total | 5152 (100) | 3620 (70.3) | 1130 (21.9) | 402 (7.8) |  | 3633 (70.5) | 1519 (29.5) |  |
| Alcohol consumption level |  |  |  |  | - |  |  | <0.0001 |
| None/low-risk | 3620 (70.3) | - | - | - |  | 2426 (66.8) | 1194 (78.6) |  |
| Moderate-risk | 1130 (21.9) | - | - | - |  | 903 (24.9) | 227 (14.9) |  |
| High-risk | 402 (7.8) | - | - | - |  | 304 (8.4) | 98 (6.5) |  |
| Physical activity |  |  |  |  | <0.0001 |  |  | - |
| Active | 3633 (70.5) | 2426 (67.0) | 903 (79.9) | 304 (75.6) |  | - | - |  |
| Sedentary | 1519 (29.5) | 1194 (33.0) | 227 (20.1) | 98 (24.4) |  | - | - |  |
| Age |  |  |  |  | <0.0001 |  |  | 0.0023 |
| 18-29 | 1129 (21.9) | 756 (20.9) | 275 (24.3) | 98 (24.4) |  | 787 (21.7) | 342 (22.5) |  |
| 30-39 | 1486 (28.8) | 1008 (27.9) | 324 (28.7) | 154 (38.3) |  | 1041 (28.7) | 445 (29.3) |  |
| 40-49 | 945 (18.3) | 686 (19.0) | 173 (15.3) | 86 (21.4) |  | 629 (17.3) | 316 (20.8) |  |
| 50-64 | 699 (13.6) | 507 (14.0) | 159 (14.1) | 33 (8.2) |  | 511 (14.1) | 188 (12.4) |  |
| 65+ | 893 (17.3) | 663 (18.3) | 199 (17.6) | 31 (7.7) |  | 665 (18.3) | 228 (15.0) |  |
| Sex |  |  |  |  | <0.0001 |  |  | <0.0001 |
| Male | 2337 (45.4) | 1647 (45.5) | 437 (38.7) | 253 (62.9) |  | 1737 (47.8) | 600 (39.5) |  |
| Female | 2815 (54.6) | 1973 (54.5) | 693 (61.3) | 149 (37.1) |  | 1896 (52.2) | 919 (60.5) |  |
| Race/ethnicity |  |  |  |  | <0.0001 |  |  | <0.0001 |
| Hispanic | 849 (16.5) | 607 (16.8) | 163 (14.4) | 79 (19.7) |  | 532 (14.6) | 317 (20.9) |  |
| White, non-Hispanic | 3225 (62.6) | 2201 (60.8) | 767 (67.9) | 257 (63.9) |  | 2404 (66.2) | 821 (54.1) |  |
| Black, non-Hispanic | 547 (10.6) | 396 (10.9) | 105 (9.3) | 46 (11.4) |  | 315 (8.7) | 232 (15.3) |  |
| Asian/Pacific Islander | 368 (7.1) | 293 (8.1) | 64 (5.7) | 11 (2.7) |  | 278 (7.7) | 90 (5.9) |  |
| Other, non-Hispanic | 163 (3.2) | 123 (3.4) | 31 (2.7) | 9 (2.2) |  | 104 (2.9) | 59 (3.9) |  |
| Education |  |  |  |  | <0.0001 |  |  | <0.0001 |
| High school or less | 609 (11.8) | 480 (13.3) | 80 (7.1) | 49 (12.2) |  | 278 (7.7) | 331 (21.8) |  |
| AIM 1 TABLES |  |  |  |  |  |  |  |  |
| Some college | 1350 (26.2) | 990 (27.4) | 251 (22.2) | 109 (27.1) |  | 791 (21.8) | 559 (36.8) |  |
| College graduate | 3193 (62.0) | 2150 (59.4) | 799 (70.7) | 244 (60.7) |  | 2564 (70.6) | 629 (41.4) |  |
| Annual household income |  |  |  |  | <0.0001 |  |  | <0.0001 |
| <\$35,000 | 1426 (27.7) | 1122 (31.0) | 198 (17.5) | 106 (26.4) | | 770 (21.2) | 656 (43.2) | |
| \$35,000-\$49,999 | 600 (11.7) | 434 (12.0) | 117 (10.4) | 49 (12.2) | | 366 (10.1) | 234 (15.4) | |
| \$50,000-\$69,999 | 771 (15.0) | 550 (15.2) | 167 (14.8) | 54 (13.4) | | 553 (15.2) | 218 (14.4) | |
| \$70,000-\$99,999 | 879 (17.1) | 602 (16.6) | 214 (18.9) | 63 (15.7) | | 679 (18.7) | 200 (13.2) | |
| \$100,000 | 1476 (28.7) | 912 (25.2) | 434 (38.4) | 130 (32.3) | | 1265 (34.8) | 211 (13.9) | |
| BMI category |  |  |  |  | <0.0001 |  |  | <0.0001 |
| Underweight | 107 (2.1) | 86 (2.4) | 17 (1.5) | 4 (1.1) |  | 74 (2.1) | 33 (2.2) |  |
| Healthy | 1873 (37.4) | 1235 (35.1) | 503 (45.7) | 135 (35.6) |  | 1492 (42.3) | 381 (25.8) |  |
| Overweight | 1505 (30.1) | 1031 (29.3) | 336 (30.5) | 138 (36.4) |  | 1125 (31.9) | 380 (25.8) |  |
| Obesity | 1519 (30.4) | 1172 (33.3) | 245 (22.3) | 102 (26.9) |  | 837 (23.7) | 682 (46.2) |  |
| In a relationship<sup>§§</sup> |  |  |  |  | <0.0001 |  |  | <0.0001 |
| Yes | 3192 (62.7) | 2158 (60.4) | 786 (70.1) | 248 (62.2) |  | 2371 (66.1) | 821 (54.5) |  |
| No | 1901 (37.3) | 1414 (39.6) | 336 (30.0) | 151 (37.8) |  | 1216 (33.9) | 685 (45.5) |  |
| Chronic health condition dx or poor general health |  |  |  |  | <0.0001 |  |  | <0.0001 |
| Yes | 2011 (38.6) | 1486 (41.1) | 366 (32.4) | 159 (39.5) |  | 1249 (34.4) | 762 (50.2) |  |
| No | 3141 (61.4) | 2134 (58.9) | 764 (67.6) | 243 (60.5) |  | 2384 (65.6) | 757 (49.8) |  |
| In recovery from drugs<sup>§§</sup> |  |  |  |  | <0.0001 |  |  | <0.0001 |
| Yes | 447 (8.9) | 313 (8.9) | 78 (7.1) | 56 (14.3) |  | 220 (6.2) | 227 (15.4) |  |
| No | 4564 (91.1) | 3210 (91.1) | 1019 (92.9) | 335 (85.7) |  | 3314 (93.8) | 1250 (84.6) |  |
| In recovery from alcohol<sup>§</sup> |  |  |  |  | <0.0001 |  |  | <0.0001 |
| Yes | 506 (9.8) | 326 (9.0) | 100 (8.9) | 80 (19.9) |  | 276 (7.6) | 230 (15.2) |  |
| No | 4645 (90.2) | 3294 (91.0) | 1029 (91.1) | 322 (80.1) |  | 3357 (92.4) | 1288 (84.9) |  |

‡Chi-squared test used for assessment of statistically significant differences across the three groups
§<1% missing §§ 1-3% missing; §§§ 4-10% missing

### Physical activity (effect modifier)

Physical activity was assessed across ten follow-up rounds from September 2021 through December 2023. Weekly physical activity was derived from a 3-item series adapted from the 2023 Behavioral Risk Factor Surveillance System, a nationally representative survey on health behaviors, chronic conditions, and preventive care among U.S. adults. Participants were asked: (1) whether they engaged in any physical activities or exercises outside of their regular job during the past month (e.g., running, calisthenics, golf, gardening, or walking for exercise) [Yes/No/Unsure]; (2) how often they engaged in these activities per week or per month; and (3) the usual duration (hours or minutes) of each session. Responses were used to calculate the average weekly minutes of physical activity over the past month. For the primary analysis, we created a binary variable for persistent activity status: participants reporting 0 minutes of activity in at least five of the ten assessments were classified as sedentary, while all others were classified as active, as we have previously done to classify persistent exposures^54,55^. Among participants classified as active, the median weekly physical activity across assessments was 177 minutes (IQR: 108-288), compared with 18 minutes (IQR: 0-49) among those classified as sedentary.

Additionally, for use in sensitivity analyses, participants were classified as consistently meeting U.S. physical activity guidelines if they reported ≥150 minutes of moderate-intensity activity per week in at least five assessments; all others were classified as not meeting guidelines.^56,57^ Among participants meeting physical activity guidelines, the median weekly physical activity across assessments was 259 minutes (IQR: 192–378), compared with 61 minutes (IQR: 18–107) among those not meeting guidelines.

### Anxiety and/or depression symptoms (outcome)

Symptoms of anxiety and depression were assessed using the validated Generalized Anxiety Disorder-7 (GAD-7; score range: 0-21) and Patient Health Questionnaire-8 (PHQ-8; score range: 0-24). Both instruments are widely used in clinical and research settings. Scores of ≥10 on either scale indicate moderate to severe symptomatology.^58–62^ To ensure the outcome was measured after exposure ascertainment, symptoms of anxiety/depression was defined as having a PHQ-8 or GAD-7 score ≥10 at the December 2023 assessment.

### Covariates

To facilitate interpretation of observed associations, we selected a limited set of baseline characteristics that may be related to both exposure and outcome, including age, gender, race/ethnicity, education, income, relationship status, recovery from alcohol or drug use, and chronic conditions or poor general health. Definitions and timing of measurement are provided in Supplementary Table S1.

### Imputation of missing data

Missing data for alcohol use, physical activity, and anxiety/depression symptoms were addressed using multiple imputation, assuming data were missing at random (MAR), and were modeled as zero-inflated continuous variables using Markov Chain Monte Carlo methods. Across the ten assessments, 72% completed all surveys and 91% completed at least six surveys (Supplementary Table S2). Ten imputations were generated for each variable. Each model included prior values of the same variable and relevant sociodemographic and health covariates. Imputed values were constrained to valid ranges, rounded to the nearest integer, and combined using weighted averages (each with weight = 0.1) to produce a single dataset for analysis, which has been shown to provide unbiased estimates of missing values.^63–65^ Model convergence and stability were verified across iterations and diagnostic checks confirmed that imputed and observed distributions were comparable.

### Statistical analysis

Proportions and chi-square tests were used to examine bivariate associations between sociodemographic characteristics, alcohol consumption, and physical activity, as well as between anxiety/depression symptoms, alcohol consumption and physical activity. Log-binomial regression models (crude and adjusted), including an interaction term, were used to estimate risk ratios (RRs) for the association between alcohol consumption and anxiety/depression symptoms, within levels of physical activity, assessing effect modification on the multiplicative scale. For interpretive consistency across analyses, physically active participants were treated as the reference group (“unexposed”), and sedentary participants as the comparison group (“exposed”), such that higher RRs indicate elevated risk among sedentary individuals. Effect modification on the additive scale was evaluated using covariate-adjusted models. The relative excess risk due to interaction (RERI) was calculated to quantify whether the combined effect of alcohol use and physical activity on anxiety/depression symptoms exceeded the sum of their individual effects. A positive RERI indicates a greater-than-additive joint effect on anxiety/depression symptoms. All models conducted in the full sample (N=5,152) and in the subset of participants who did not report any chronic conditions or poor general health at enrollment (N=3,141) to reduce potential bias related to ”sick-quitters”.^66^ Analyses were conducted using SAS version 9.4 (SAS Institute, Cary, NC, USA), with statistical significance set at *p* < 0.05.

### Sensitivity Analyses

As a sensitivity analysis, all models were repeated using a binary classification of physical activity-meeting versus not meeting U.S. recommendations of ≥150 minutes per week-to evaluate the robustness of findings to different operationalizations of physical activity.

## Results

### Sample characteristics by alcohol consumption and physical activity level

Characteristics for the analytic sample are shown in Table 1. This full cohort consisted of 5,152 participants. Most (70.3%) reported low-risk or no alcohol use, 21.9% reported moderate-risk use and 7.8% reported high-risk use. Overall, 70.5% were classified as active (versus sedentary), and 40.8% met physical activity guidelines of 150+ minutes per week.

Non/low-risk and high-risk drinkers shared several characteristics that distinguished them from moderate-risk drinkers (Table 1). They were less likely to have graduated from college (59.4%–60.7% vs. 70.7%), to be White, non-Hispanic (60.8-63.9% vs. 67.9%), to be in a relationship (60.4-62.2% vs. 70.1%) and to report household incomes ≥$100,000 (25.2–32.3% vs. 38.4%). They were also more likely to have chronic conditions or poor general health (39.5–41.1% vs. 32.4%). Compared with both moderate and non/low-risk drinkers, high-risk drinkers were more likely to be male (62.9% vs. 38.7– 45.5%), aged 30–39 (38.3% vs. 27.9–28.7%), and to report being in recovery from drugs (14.3% vs. 7.1-8.9%) or alcohol (19.9% vs. 8.9-9.0%), and were less likely to identify as Asian/Pacific Islander (2.7% vs. 5.7–8.1%).

Greater alcohol consumption was associated with greater physical activity, with both moderate and high-risk drinkers more likely than non/low-risk drinkers to be physically active (vs. sedentary) (75.6%-79.9% vs. 67.0%). One-third (33.3%) of physically active participants were moderate or high-risk drinkers, compared to 21.4% of sedentary participants. Compared with sedentary participants, those who were physically active were more likely to be male (47.8% vs. 39.5%), White non-Hispanic (66.2% vs. 54.1%), college graduates (70.6% vs. 41.4%), and to report household incomes ≥$100,000 (34.8% vs. 13.9%). Active participants were also more likely to have a healthy body mass index (42.3% vs. 25.8%) and to be in a relationship (66.1% vs. 54.5%). In contrast, they were less likely to report chronic conditions or poor general health (34.4% vs. 50.2%) and less likely to report being in recovery from drugs or alcohol (10.7% vs. 22.5%).

Sociodemographic patterns were generally similar in the restricted sample without chronic conditions or poor health (Supplementary Table S3).

### Prevalence of anxiety/depression symptoms by alcohol use and physical activity

Overall, 21.7% of participants met criteria for moderate to severe anxiety/depression symptoms in December 2023 (Table 2). Of these, 54.5% had symptoms of both conditions, 30.1% had symptoms of depression only, and 15.4% had symptoms of anxiety only. The prevalence of anxiety/depression symptoms differed significantly across alcohol and physical activity categories. Participants with high-risk alcohol use had the highest prevalence of anxiety/depression symptoms (28.4%) compared with those with no/low-risk (21.5%) and those with moderate-risk (20.0%). Anxiety/depression symptoms were less common among physically active participants (16.5%) compared to sedentary (34.2%). In the subsample without underlying health conditions, the overall prevalence of anxiety/depression symptoms was 18.9% and followed a similar pattern by physical activity level: 15.9% among active versus 28.3% among sedentary participants. Differences across alcohol use categories were smaller and not statistically significant (*p* = 0.22).

**Table 2.**
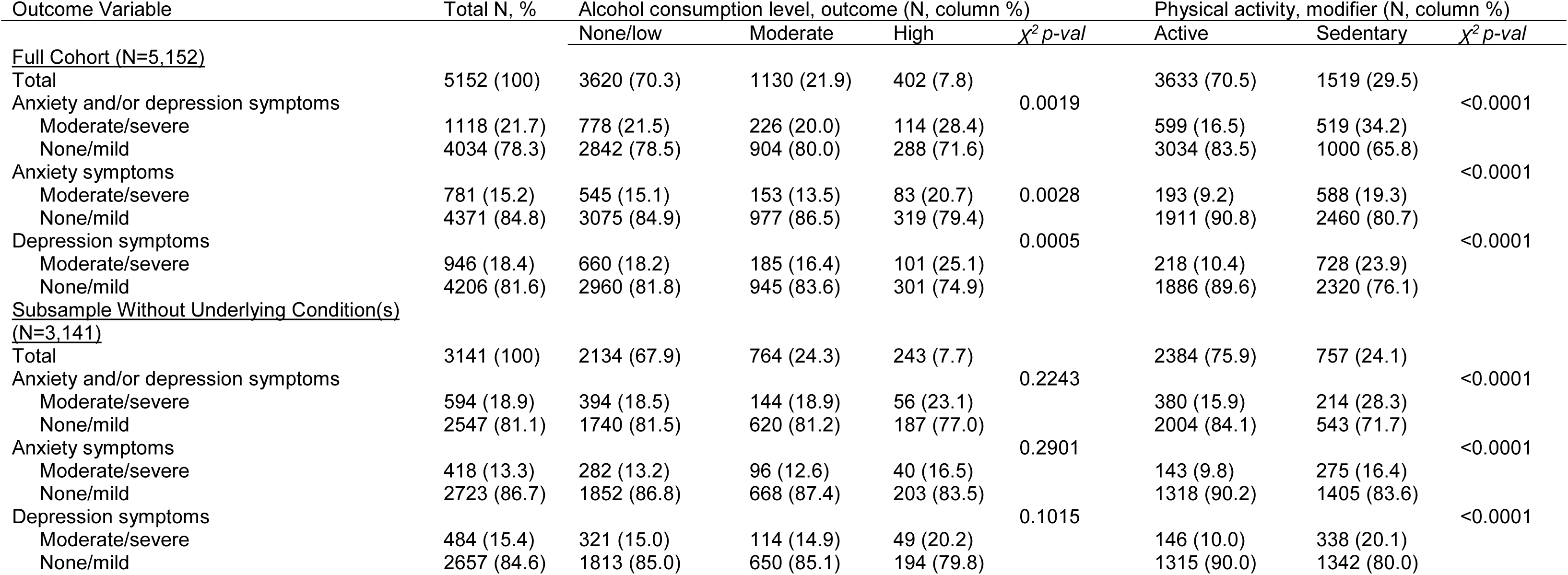
Anxiety and/or depression symptoms (outcome) by alcohol consumption level (exposure) and physical activity level (modifier), CHASING COVID Cohort, September 2021-December 2023.

### Effect measure modification

For the full sample, the highest risk of anxiety/depression symptoms was observed among sedentary participants with high-risk alcohol use compared with active participants with low-risk or no alcohol use (aRR=1.96, 95% CI: 1.54-2.48) (Table 3). Within physical activity strata, moderate-risk alcohol use was not associated with anxiety/depression symptoms, while high-risk alcohol use was associated with elevated risk only among active participants. The relative risk of anxiety/depression symptoms for high-risk drinkers compared with non/low-risk drinkers was1.44 (95% CI: 1.15-1.80) among active participants and 1.17 (95% CI: 0.93-1.46), among sedentary participants. There was no evidence of effect modification by physical activity on either the additive or multiplicative scales. The RERI was 0.04 (95% CI: −0.23, 0.31) for moderate drinking and −0.24 (95% CI: −0.68, 0.19) for high-risk drinking. Multiplicative interaction estimates were similarly null (moderate: 1.04, 95% CI: 0.81-1.34; high-risk: 0.81, 95% CI: 0.59-1.11).

**Table 3.** Effect modification for the association between alcohol consumption risk level and anxiety or depression symptoms within levels of physical activity (active vs. sedentary), CHASING COVID Cohort, September 2021-December 2023 (N=5,152)

|  |  |  |  |  |  |
| --- | --- | --- | --- | --- | --- |
|  |  | Active |  | Sedentary |  |
|  |  | *N* with/without moderate or severe symptoms of anxiety/depression | RR (95% CI); *P* | *N* with/without moderate or severe symptoms of anxiety/depression | RR (95% CI); *P* |
| Alcohol consumption risk level | None/Low | 384/2042 | 1.00 (reference) | 394/800 | 1.68 (1.47, 1.90) |
|  | Moderate | 142/761 | 0.98 (0.82, 1.17) | 84/143 | 1.70 (1.41, 2.06) |
|  | High | 73/231 | 1.44 (1.15, 1.80) | 41/57 | 1.96 (1.54, 2.48) |
| RRs (95% CI) for moderate-risk alcohol consumption within strata of physical activity |  |  | 0.98 (0.82, 1.17) |  | 1.02 (0.85, 1.21) |
| RRs (95% CI) for high-risk alcohol consumption within strata of physical activity |  |  | 1.44 (1.15, 1.80) |  | 1.17 (0.93, 1.46) |
| Measure of interaction on additive scale (moderate): |  |  | 0.04 (-0.23, 0.31), p=0.7676 |  |  |
| Measure of interaction on additive scale (high): |  |  | -0.24 (-0.68, 0.19), p=0.2749 |  |  |
| Measure of interaction on multiplicative scale (moderate): |  |  | 1.04 (0.81, 1.34), p=0.7651 |  |  |
| Measure of interaction on multiplicative scale (high): |  |  | 0.81 (0.59, 1.11), p=0.1956 |  |  |
| RRs adjusted for age, gender, race/ethnicity, education, body mass index, relationship status, or any chronic conditions |  |  |  |  |  |

Among participants without underlying health conditions, patterns of association differed from those observed in the full sample, with evidence of effect modification by physical activity (Table 4). The highest risk of anxiety/depression symptoms was observed among sedentary participants with moderate-risk alcohol use, who had nearly twice the risk compared with active participants with low-risk or no alcohol use (aRR=1.92, 95% CI: 1.50-2.45). Sedentary high-risk drinkers had elevated risk (aRR=1.62, 95% CI: 1.03-2.55) compared with low-no risk active participants and sedentary non/low-risk drinkers had higher risk than low-no risk active participants (aRR=1.36, 95% CI: 1.12-1.63). Within physical-activity strata, the relative risk of anxiety/depression symptoms for moderate drinkers compared with non/low-risk drinkers was 0.95 (95% CI: 0.76-1.19) among active participants and 1.41 (95% CI: 1.10-1.82) among sedentary participants. For high-risk drinkers, the corresponding aRRs were 1.29 (95% CI: 0.95-1.75) and 1.20 (95% CI: 0.76-1.88), respectively. Evidence of effect modification by physical activity was detected for moderate drinkers on both the additive and multiplicative scales (RERI = 0.61, 95% CI: 0.13-1.09; interaction aRR=1.49, 95% CI: 1.07-2.07), indicating that the joint effect of moderate alcohol use and sedentary behavior on anxiety/depression symptoms exceeded the sum of their individual effects. No significant interaction was observed for high-risk drinking on either scale.

**Table 4.** Effect modification of the association between alcohol consumption risk and anxiety/depression symptoms by physical activity level (active vs. sedentary) among participants without chronic conditions or poor health, CHASING COVID Cohort, September 2021–December 2023 (N=3,141)

|  |  |  |  |  |  |
| --- | --- | --- | --- | --- | --- |
|  |  | Active |  | Sedentary |  |
|  |  | *N* with/without moderate to severe anxiety/depression symptoms | RR (95% CI); *P* | *N* with/without moderate to severe anxiety/depression symptoms | RR (95% CI); *P* |
| Alcohol consumption risk level | None/Low | 243/1315 | 1.00 (reference) | 151/425 | 1.36 (1.12, 1.63) |
|  | Moderate | 96/536 | 0.95 (0.76, 1.19) | 48/84 | 1.92 (1.50, 2.45) |
|  | High | 41/153 | 1.29 (0.95, 1.75) | 15/34 | 1.62 (1.03, 2.55) |
| RRs (95% CI) for moderate-risk alcohol consumption within strata of physical activity |  |  | 0.95 (0.76, 1.19) |  | 1.41 (1.10, 1.82) |
| RRs (95% CI) for high-risk alcohol consumption within strata of physical activity |  |  | 1.29 (0.95, 1.75) |  | 1.20 (0.76, 1.88) |
| Measure of interaction on additive scale (moderate): |  |  | **0.61 (0.13, 1.09), p=0.0129** |  |  |
| Measure of interaction on additive scale (high): |  |  | -0.02 (-0.84, 0.80), p=0.9660 |  |  |
| Measure of interaction on multiplicative scale (moderate): |  |  | **1.49 (1.07, 2.07), p=0.0014** |  |  |
| Measure of interaction on multiplicative scale (high): |  |  | 0.93 (0.54, 1.61), p=0.7988 |  |  |
| RRs adjusted for age, gender, race/ethnicity, education, body mass index, relationship status |  |  |  |  |  |

### Sensitivity analysis

In the full sample, results were consistent when physical activity was defined according to whether participants met guidelines of at least 150 minutes per week (Supplementary Table S4). Within physical activity strata, the relative risk of anxiety/depression symptoms for moderate drinkers compared with non/low-risk drinkers was 0.97 (95% CI: 0.74-1.26) among those meeting guidelines and 1.01 (95% CI: 0.87-1.17) among those not meeting guidelines. For high-risk drinkers, the corresponding aRRs were 1.53 (95% CI: 1.12-2.10) and 1.23 (95% CI: 1.02-1.47), respectively. There was no statistically significant evidence of effect modification by meeting activity guidelines on either the additive or multiplicative scales (RERI = 0.05, 95% CI: −0.30, 0.40; interaction aRR=1.05, 95% CI: 0.77-1.41 for moderate drinking).

In contrast, among participants without chronic conditions or poor general health, results differed from the primary analysis (Supplementary Table S5). Within physical activity strata, moderate drinking was not associated with anxiety/depression symptoms among those meeting guidelines (RR = 0.97, 95% CI: 0.71-1.32) or among those not meeting guidelines (RR = 1.17, 95% CI: 0.96-1.44). Similarly, no interaction was observed for high-risk drinking (RERI = 0.27, 95% CI: −0.15, 0.69; interaction RR = 1.21, 95% CI: 0.84-1.74).

## Discussion

In this longitudinal cohort of U.S. adults, physical activity modified the association between alcohol use and symptoms of anxiety or depression among individuals without underlying health conditions, a subgroup less susceptible to “sick-quitter” bias. On the multiplicative scale, moderate alcohol use was associated with significantly greater risk of anxiety and depression symptoms among sedentary participants (aRR=1.41, 95% CI: 1.10-1.82) but not among active participants (aRR=0.95, 95% CI: 0.76-1.19) (interaction aRR=1.49, p<0.01), highlighting physical activity as an important contextual factor influencing alcohol-mental health associations and suggesting that physical activity may be beneficial for mitigating adverse mental health correlates of moderate alcohol consumption among healthy adults. On the additive scale, the combined presence of moderate drinking and sedentary behavior was associated with greater anxiety or depression symptom risk than would be expected based on the sum of their individual associations (RERI=0.61, p<0.01), suggesting an amplified anxiety and depression symptom burden among sedentary moderate-risk drinkers. Findings indicate that screening approaches that jointly consider alcohol use and physical activity may increase identification of individuals at increased risk of anxiety and depression symptom burden.

Using an alternative definition of physical activity based on guideline thresholds (meeting vs. not meeting ≥150 minutes per week) yielded no evidence of effect modification, suggesting that the absence of physical activity, rather than failure to meet specific guideline targets, may shape how alcohol use relates to symptoms of anxiety and depression. While we found no evidence of effect modification in the full sample, this is likely due to inflated risk among abstainers which can weaken associations. Notably, interaction was not observed for the sedentary high-risk drinkers, but the small size of this subgroup resulted in imprecise estimates limiting our ability to characterize modification at this level of exposure. Participants classified as sedentary in the primary analysis reported minimal average activity across all assessments (median 18 minutes per week, IQR: 0-49), whereas those classified as active reported substantially higher levels (median 177 minutes per week, IQR: 108-288), indicating that the comparison reflects near absence of activity versus regular, near-guideline activity levels. Future research should clarify the physical activity levels at which modification becomes apparent, ideally in cohorts with larger numbers of high-risk drinkers to improve precision in these strata.

Sedentary adults had higher risk of anxiety/depression symptoms than active non/low-risk drinkers at every alcohol consumption level, both in the full sample (aRRs 1.68-1.96) and in the subsample without underlying conditions (aRRs 1.36-1.92). In contrast, among active adults, elevated risk of anxiety/depression symptoms was found only among high-risk drinkers relative to low-no risk. This pattern suggests that physical activity may play a larger role than alcohol consumption in shaping the symptom burden of depression and anxiety, potentially obscuring alcohol-mental health associations in studies that do not adjust for activity patterns. Notably, the role of physical activity has often been overlooked in alcohol-mental health research, despite the fact adults with high physical activity are more likely to consume alcohol moderately or heavily compared to lower activity counterparts.^27–31^ A 2019 systematic review and meta-analysis identified 42 studies examining the association between alcohol use and depressive symptoms. Only ten controlled for physical activity, and nine of these identified null associations for moderate and heavy intake compared with abstainers.^40^ Together with this literature, our findings suggest that unmeasured confounding by physical activity may contribute to inconsistent findings and may partly underlie the appearance of J-shaped associations in prior studies.

### Strengths and limitations

This study leveraged a large and socio-demographically diverse U.S. cohort with repeated measures of alcohol use, physical activity, and symptoms of anxiety or depression over two years, enabling examination of behavioral and mental health patterns across diverse social and economic contexts.

Repeated assessments also allowed us to incorporate duration of exposure into the alcohol and physical activity measures and to ensure that exposures preceded outcomes in time. However, several limitations should be noted. All measures were self-reported and may be subject to misclassification.

Symptoms of anxiety and depression were assessed using screening instruments rather than clinical diagnoses; therefore, findings should be interpreted as pertaining to symptom burden, not diagnosed disorders. We also examined anxiety or depression symptoms as a combined outcome and did not evaluate whether associations differed for anxiety symptoms, depressive symptoms, or co-occurring symptoms, which may have obscured heterogeneity across mental health symptom profiles. Given the limited set of adjustment variables and inability to control for time-varying confounding, findings should be interpreted as descriptive rather than causal. We also did not incorporate receipt of mental health treatment, which may shape observed associations. Multiple imputation relied on the assumption that data were missing at random conditional on variables included in the imputation models, which may not fully hold in practice. Subgroup analyses were additionally limited by small cell sizes, particularly among sedentary high-risk drinkers, yielding imprecise estimates and wide confidence intervals likely due to limited statistical power to detect associations. Finally, while the cohort was diverse, it was not nationally representative and participants were recruited into a COVID-19-focused study, which may limit generalizability if individuals who enroll in pandemic-related research differ systematically from the U.S. population.

Together, these findings suggest that, in the absence of health-related behavioral constraints, even modest physical activity may help offset some of the burden of depression and anxiety symptoms associated with greater alcohol use. They also highlight the importance of jointly considering alcohol consumption and movement patterns when identifying groups at elevated mental health risk. Future prospective and intervention studies using causal inference approaches are warranted to further clarify these relationships and inform interventions targeting modifiable lifestyle-related risk factors.

## Data Availability

The datasets generated and/or analyzed during the current study are available in the repository, Zenodo: DOI: 10.5281/zenodo.6127734. Some data elements are not publicly available due to funder requirements, but are available from the authors upon reasonable request, subject to approval and available resources.

https://zenodo.org/records/7305435

## Acknowledgements

We thank the participants of the Communities, Households, and SARS-CoV-2 Epidemiology COVID Cohort Study for their contribution to the advancement of science.

## CRediT Author Statement

**Jenna Sanborn:** Conceptualization, Methodology, Software, Formal analysis, Investigation, Writing-Original Draft. **Denis Nash**: Writing-Review & Editing, Funding acquisition. **McKaylee Robertson:** Writing-Review & Editing. **Angela M Parcesepe**: Writing-Review & Editing, Funding acquisition. **Zachary Shahn:** Methodology, Writing-Review and Editing, Supervision.

## Data availability statement

The datasets generated and/or analyzed during the current study are available in the repository, Zenodo: <u>DOI: 10.5281/zenodo.6127734</u>. Some data elements are not publicly available due to funder requirements, but are available from the authors upon reasonable request, subject to approval and available resources.

