## Supplementary Material for "The role of physical activity in shaping the relationship between alcohol use and anxiety and depression symptoms: Findings from a U.S. nationwide cohort"

| **Supplemental Table S1. Definitions and assessment timing of variables used in analyses** | | | | |
| --- | --- | --- | --- | --- |
| **Variable** | **Variable type** | **Assessment Timing** | **Definition** | **Survey Question(s)** |
| Depression and/or anxiety symptoms (outcome) | Binary | Dec 2023 | Coded as ‘Yes’ if moderate or severe symptoms endorsed in any assessment, otherwise ‘No’ | The GAD-7 assesses anxiety symptoms, and the PHQ-8 assesses depression symptoms.  GAD-7 includes seven items asking how often participants experienced specific symptoms over the past two weeks. Response options range from 0 = not at all to 3 = nearly every day, yielding a total score from 0 to 21. Higher scores indicate greater symptom severity. Consistent with standard practice, scores ≥10 were used to identify moderate-to-severe anxiety symptom. Items included: 1. Feeling nervous, anxious or on edge 2. Not being able to stop or control worrying 3. Worrying too much about different things 4. Trouble relaxing 5. Being so restless that it’s hard to sit still 6. Being easily annoyed or irritable 7. Feeling afraid as if something awful might happen  PHQ-8 includes eight items asking how often participants experienced specific symptoms over the past two weeks. Response options range from 0 = not at all to 3 = nearly every day, yielding a total score from 0 to 24. Higher scores indicate greater symptom severity. Consistent with standard practice, scores ≥10 were used to identify moderate-to-severe depressive symptom. Items included: 1. Little interest or pleasure in doing things 2. Feeling down, depressed or hopeless 3. Trouble falling or staying asleep, or sleeping too much 4. Feeling tired or having little energy 5. Poor appetite or overeating 6. Feeling bad about yourself - or that you are a failure or have let yourself or your family down 7. Trouble concentrating on things, such as, reading the newspaper or watching television 8. Moving or speaking so slowly that other people could have noticed? Or the opposite - being so fidgety or restless that you have been moving around a lot more than usual. |
| Alcohol consumption level (exposure) | Categorical | Sept 2021, Mar 2022, Oct 2022, Apr 2023, Sept 2023 | Scores were categorized using sex-specific thresholds: for males, 0–3 = low-risk, 4–5 = moderate-risk, ≥6 = high-risk; for females, 0–2 = low-risk, 3–5 = moderate-risk, ≥6 = high-risk.  Participants were classified hierarchically: Those meeting criteria for high-risk drinking in ≥50% of assessments were classified as persistently high-risk; among the remainder, those with low-risk drinking in ≥50% of assessments were classified as persistently low-risk; all others were classified as moderate-risk. | In the last month, how often do you have a drink containing alcohol? a) Never b) Monthly or less c) 2-4 times a month d) 2-3 times a week e) 4 or more times a week (scored 0 to 4)  If never drank alcohol in the last month, then skip: In the last month, how many standard drinks containing alcohol do you have on a typical day? One standard drink is equal to 12 ounces of beer, 5 ounces of wine, 1.5 ounces of hard liquor or 8 ounces of malt liquor. a) 1 or 2 b) 3 or 4 c) 5 or 6 d) 7 or 9 e) 10 or more (Scored 0-4)  f never drank alcohol in the last month, then skip: In the last month, how often do you have six or more drinks on one occasion? a) Never b) Less than monthly (Score 0-4) |
| Physical activity (modifier) | Binary | Sept 2021, Dec 2021, Mar 2022, Jun 2022, Oct 2022, Dec 2022, Apr 2023, Jun 2023, Sept 2023, Dec 2023 | Minutes per week derived from item responses. Two binary classifications were obtained:  1) Sedentary vs. active. ‘Sedentary’ defined as having 0 minutes of exercise per week in at least 50% of assessments, otherwise ‘active’; and  2) Meeting recommendations vs. not: exercising 150+ minutes per week vs. <150 minutes per week | During the past month, other than your regular job, did you participate in any physical activities or exercises such as running, calisthenics, golf, gardening, or walking for exercise? a) Yes b) No c) Don’t know / Not sure  How many times per week or per month did you take part in this activity during the past month? a) __ Times per week b) __ Times per month c) Don’t know / Not sure  And when you took part in this activity, for how many minutes or hours did you usually keep at it? a) __ Number of hours b) __ Number of minutes c) Don’t know / Not sure |
| Sociodemographics |  |  |  |  |
| Age | Continuous | Enrollment |  | What is your age? ____ |
| Sex | Binary | April 2023 | Collapsed to 2 categories: ‘Male’ and ‘Female’. Missing values (n=198), imputed with gender identity, assessed at enrollment (if male or female endorsed) | What sex were you assigned at birth? a) Male, b) Female, c) None of the above |
| Race/Ethnicity | Categorical | Enrollment | Collapsed to five categories: 1) Hispanic, 2) White, non-Hispanic, 3) Black, non-Hispanic, 4) Asian, non-Hispanic and 5) Other, non-Hispanic | Are you Hispanic, Latino/a, or Spanish origin? a) Yes b) No c) Don’t know / Not sure  Which of these groups would you say best represents your race? Please select all that apply. a) Black or African American, b) American Indian or Alaska Native, c) Asian, d) Pacific Islander, e) White, f) Other____ |
| Education | Categorical | Enrollment | Collapsed to three categories: 1) High school or less, 2) Some college, and 3) College graduate | What is the highest grade or year of school you completed? a) Less than a high school diploma, b) Grade 12 or GED (High school graduate), c) College 1 year to 3 years (Some college or technical school), d) College 4 years or more (College graduate) |
| Annual household income | Categorical | Enrollment | Collapsed to three categories: 1) <49,000, 2) $50,000-$99,999 and 3) $100,000+ | Is your annual household income from all sources a) <$25,000, b) $25,000-$34,999, c) $35,000-$49,999, d) $50,000-$69,999, e) $70,000-$99,999, f) $100,00-$149,000, g) $150,000+ |
| Body Mass Index | Categorical | Enrollment | Calculated as kg/m^2^ based on height and weight, categorized as Normal, Healthy, Overweight and Obesity based on guidelines from the centers for disease control and prevention (CDC) | How much do you weigh without shoes? Please answer in pounds ____  About how tall are you without shoes? Please answer in feet and inches. ____ Feet ____ Inches |
| Relationship status | Binary | Sept 2021, May 2021, Feb 2021, Dec 2020, Nov 2020, Oct 2020, July 2020, Enrollment  Most recent non-missing value used | To categories: 1) Yes, 2) No/Don’t know | Are you currently in a relationship or seeing someone? a) Yes b) No c) Don’t know/ Not sure |
| Chronic health condition diagnosis or poor general health status | Binary | Enrollment | If any chronic conditions are selected, or if poor general health is endorsed, then categorized as ‘Yes’; otherwise categorized as ‘No’ | Has a doctor, nurse, or other health professional ever told you that you had any of the following? Please select all that apply a) had a heart attack also called a myocardial infarction? b) have angina or coronary heart disease? c) have type 2 diabetes? d) have high blood pressure? e) have cancer? f) had asthma? g) have chronic obstructive pulmonary disease, C.O.P.D., emphysema or chronic bronchitis? h) have kidney disease (not including kidney stones, bladder infection or incontinence)? i) have HIV/AIDS? j) have immunosuppression? k) have chronic liver disease, including cirrhosis?  Would you say that in general your health is: a) Excellent b) Very good c) Fair d) Poor |
| Ever in recovery for drugs or alcohol | Binary | Drug recovery: Sept 2021, May 2021, Dec 2020, July 2020  Alcohol recovery: Sept 2021, May 2021, Nov 2020, Enrollment | If ‘I am currently in recovery’ or ‘I am not currently in recovery, but have been previously’ are endorsed for either question at any timepoint, then participant categorized as having ever been in recovery from drugs or alcohol, otherwise not. | Do you identify as being in recovery from drugs? a) Yes, I am currently in recovery from drugs, b) I am not currently in recovery from drugs, but I have previously been in recovery from drugs, c) I have never been in recovery from drugs  Do you identify as being in recovery from alcohol? a) Yes, I am currently in recovery from alcohol, b) I am not currently in recovery from alcohol, but I have previously been in recovery from alcohol, c) I have never been in recovery from alcohol, d) Don’t know/ Not sure |

Supplementary Table S2. Number and percentage of

participants by number of missing assessments

(Sept 2021- Dec 2023)

| Number of missing assessments | N | % |
| --- | --- | --- |
| 0 | 3,724 | 72.3 |
| 1 | 472 | 9.2 |
| 2 | 194 | 3.8 |
| 3 | 140 | 2.7 |
| 4 | 134 | 2.6 |
| 5 | 119 | 2.3 |
| 6 | 98 | 1.9 |
| 7 | 92 | 1.8 |
| 8 | 102 | 2.0 |
| 9 | 76 | 1.5 |

| **Supplementary Table S3. Cohort characteristics by exposure and modifier status for the subsample without underlying condition(s), CHASING COVID Cohort, September 2021-December 2023 (N=3,141)** | | | | | | | | | | | |
| --- | --- | --- | --- | --- | --- | --- | --- | --- | --- | --- | --- |
|  | Total | Alcohol consumption level | | | ꭕ^2^ p-value | Physical activity | | ꭕ^2^ p-value | Physical activity | |  |
|  |  | None/low-risk | Moderate | High risk |  | Active | Sedentary |  | ≥ 150 mins/wk | < 150 mins/wk | ꭕ^2^ p-value |
|  | N (%) | n (col %) | n (col %) | n (col %) |  | n (col %) | n (col %) |  | n (col %) | n (col %) |  |
| Total | 3141 (100) | 2134 (67.9) | 764 (24.3) | 243 (7.7) |  | 2384 (75.9) | 757 (24.1) |  | 1461 (46.5) | 1680 (53.5) |  |
| Alcohol consumption level |  |  |  |  | - |  |  | <0.0001 |  |  | <0.0001 |
| None/low-risk | 2134 (67.9) | - | - | - |  | 1558 (65.3) | 576 (76.1) |  | 914 (62.6) | 1220 (72.6) |  |
| Moderate-risk | 764 (24.3) | - | - | - |  | 632 (26.5) | 132 (17.4) |  | 426 (29.2) | 338 (20.1) |  |
| High-risk | 243 (7.7) | - | - | - |  | 194 (8.1) | 49 (6.5) |  | 121 (8.3) | 122 (7.3) |  |
| Physical activity |  |  |  |  | <0.0001 |  |  | - | - | - | - |
| ≥ 150 mins/wk | 1461 (46.5) | 914 (42.8) | 426 (55.8) | 121 (49.8) |  | - | - |  | - | - |  |
| < 150 mins/wk | 1680 (53.5) | 1220 (57.2) | 338 (44.2) | 122 (50.2) |  | - | - |  | - | - |  |
| Physical activity |  |  |  |  | <0.0001 |  |  |  |  |  |  |
| Active | 2384 (75.9) | 1558 (73.0) | 632 (82.7) | 194 (79.8) |  | - | - |  | - | - |  |
| Sedentary | 757 (24.1) | 576 (27.0) | 132 (17.3) | 49 (20.2) |  | - | - |  | - | - |  |
| Age |  |  |  |  | 0.0072 |  |  | <0.0001 |  |  | <0.0001 |
| 18-29 | 858 (27.3) | 581 (27.2) | 209 (27.4) | 68 (28.0) |  | 617 (25.9) | 241 (31.8) |  | 322 (22.0) | 536 (31.9) |  |
| 30-39 | 1020 (32.5) | 679 (31.8) | 241 (31.5) | 100 (41.2) |  | 773 (32.4) | 247 (32.6) |  | 455 (31.1) | 565 (33.6) |  |
| 40-49 | 563 (17.9) | 386 (18.1) | 130 (17.0) | 47 (19.3) |  | 401 (16.8) | 162 (21.4) |  | 260 (17.8) | 303 (18.0) |  |
| 50-64 | 348 (11.1) | 242 (11.3) | 91 (11.9) | 15 (6.2) |  | 292 (12.3) | 56 (7.4) |  | 190 (13.0) | 158 (9.4) |  |
| 65+ | 352 (11.2) | 246 (11.5) | 93 (12.2) | 13 (5.4) |  | 301 (12.6) | 51 (6.7) |  | 234 (16.0) | 118 (7.0) |  |
| Sex |  |  |  |  | <0.0001 |  |  | <0.0001 |  |  | <0.0001 |
| Male | 1344 (42.8) | 929 (43.5) | 264 (34.6) | 151 (62.1) |  | 1068 (44.8) | 276 (36.5) |  | 709 (48.5) | 635 (37.8) |  |
| Female | 1797 (57.2) | 1205 (56.5) | 500 (65.5) | 92 (37.9) |  | 1316 (55.2) | 481 (63.5) |  | 752 (51.5) | 1045 (62.2) |  |
| Race/ethnicity |  |  |  |  | <0.0001 |  |  | <0.0001 |  |  | <0.0001 |
| Hispanic | 547 (17.4) | 402 (18.8) | 104 (13.6) | 41 (16.9) |  | 375 (15.7) | 172 (22.7) |  | 225 (15.4) | 322 (19.2) |  |
| White, non-Hispanic | 1925 (61.3) | 1234 (57.8) | 528 (69.1) | 163 (67.1) |  | 1545 (64.8) | 380 (50.2) |  | 964 (66.0) | 961 (57.2) |  |
| Black, non-Hispanic | 296 (9.4) | 208 (9.8) | 64 (8.4) | 24 (9.9) |  | 180 (7.6) | 116 (15.3) |  | 97 (6.6) | 199 (11.9) |  |
| Asian/Pacific Islander | 285 (9.1) | 226 (10.6) | 49 (6.4) | 10 (4.1) |  | 219 (9.2) | 66 (8.7) |  | 133 (9.1) | 152 (9.1) |  |
| Other, non-Hispanic | 88 (2.8) | 64 (3.0) | 19 (2.5) | 5 (2.1) |  | 65 (2.7) | 23 (3.0) |  | 42 (2.9) | 46 (2.7) |  |
| Education |  |  |  |  | <0.0001 |  |  | <0.0001 |  |  | <0.0001 |
| High school or less | 340 (10.8) | 267 (12.5) | 47 (6.2) | 26 (10.7) |  | 165 (6.9) | 175 (23.1) |  | 111 (5.3) | 498 (16.3) |  |
| Some college | 746 (23.8) | 522 (24.5) | 161 (21.1) | 63 (25.9) |  | 475 (19.9) | 271 (35.8) |  | 390 (18.5) | 960 (31.5) |  |
| College graduate | 2055 (65.4) | 1345 (63.0) | 556 (72.8) | 154 (63.4) |  | 1744 (73.2) | 311 (41.1) |  | 1603 (76.2) | 1590 (52.2) |  |
| Annual household income |  |  |  |  | <0.0001 |  |  | <0.0001 |  |  | <0.0001 |
| <$35,000 | 781 (24.9) | 606 (38.4) | 128 (16.8) | 47 (19.3) |  | 471 (19.8) | 310 (41.0) |  | 234 (16.0) | 547 (32.6) |  |
| $35,000-$49,999 | 340 (10.8) | 235 (11.0) | 78 (10.2) | 27 (11.1) |  | 227 (9.5) | 113 (14.9) |  | 127 (8.7) | 213 (12.7) |  |
| $50,000-$69,999 | 462 (14.7) | 327 (15.3) | 99 (13.0) | 36 (14.8) |  | 360 (15.1) | 102 (13.5) |  | 221 (15.1) | 241 (14.4) |  |
| $70,000-$99,999 | 552 (17.6) | 365 (17.1) | 149 (19.5) | 38 (15.6) |  | 448 (18.8) | 104 (13.7) |  | 278 (19.0) | 274 (16.3) |  |
| $100,000 | 1006 (32.0) | 601 (28.2) | 310 (40.6) | 95 (39.1) |  | 878 (36.8) | 128 (16.9) |  | 601 (41.1) | 405 (24.1) |  |
| BMI category |  |  |  |  | <0.0001 |  |  | <0.0001 |  |  | <0.0001 |
| Underweight | 79 (2.6) | 67 (3.2) | 10 (1.3) | 4 (0.9) |  | 57 (2.5) | 22 (3.0) |  | 30 (2.1) | 49 (3.0) |  |
| Healthy | 1392 (45.6) | 896 (43.3) | 394 (52.5) | 102 (44.2) |  | 1136 (49.1) | 256 (34.8) |  | 727 (51.2) | 665 (40.8) |  |
| Overweight | 921 (30.2) | 619 (29.9) | 217 (28.9) | 85 (36.8) |  | 715 (30.9) | 206 (28.0) |  | 449 (31.6) | 472 (28.9) |  |
| Obesity | 658 (21.6) | 487 (23.5) | 129 (17.2) | 42 (18.2) |  | 407 (17.6) | 251 (34.2) |  | 213 (15.0) | 445 (27.3) |  |
| In a relationship^§§^ |  |  |  |  | 0.0008 |  |  | <0.0001 |  |  | 0.0025 |
| Yes | 2032 (65.4) | 1343 (63.6) | 538 (71.1) | 151 (62.9) |  | 1594 (67.7) | 438 (58.2) |  | 989 (68.2) | 1043 (63.0) |  |
| No | 1075 (34.6) | 767 (36.4) | 219 (28.9) | 89 (37.1) |  | 760 (32.3) | 315 (41.8) |  | 462 (31.8) | 613 (37.0) |  |
| Ever in recovery from drugs^§§^ |  |  |  |  | 0.0005 |  |  | <0.0001 |  |  | <0.0001 |
| Yes | 223 (7.3) | 158 (7.6) | 36 (4.8) | 29 (12.2) |  | 115 (4.9) | 108 (14.7) |  | 59 (4.1) | 164 (10.1) |  |
| No | 2844 (92.7) | 1927 (92.4) | 708 (95.2) | 209 (87.8) |  | 2218 (95.1) | 626 (85.3) |  | 1383 (95.9) | 1461 (89.9) |  |
| Ever in recovery from alcohol^§^ |  |  |  |  | <0.0001 |  |  | <0.0001 |  |  | <0.0001 |
| Yes | 272 (8.7) | 169 (7.9) | 58 (7.6) | 45 (18.5) |  | 158 (6.6) | 114 (15.1) |  | 88 (6.0) | 184 (11.0) |  |
| No | 2868 (91.3) | 1965 (92.1) | 705 (92.4) | 198 (81.5) |  | 2226 (93.4) | 642 (84.9) |  | 1373 (94.0) | 1495 (89.0) |  |

‡Chi-squared test used for assessment of statistically significant differences across the three groups
^§^<1% missing ^§§^ 1-3% missing; ^§§§^ 4-10% missing

| **Supplementary Table S4. Effect modification for the association between alcohol consumption risk level and anxiety and depression within levels of physical activity (≥150 mins/week vs. <150 mins/week), CHASING COVID Cohort, September 2021-December 2023 (N=5,152)** | | | | | | |
| --- | --- | --- | --- | --- | --- | --- |
|  |  | Meets physical activity recommendations  (≥ 150 mins/week) | |  | Does not meet physical activity recommendations (< 150 mins/week) | |
|  |  | *N* with/without anxiety/depression | RR (95% CI); *P* |  | *N* with/without anxiety/depression | RR (95% CI); *P* |
| Alcohol consumption risk level | None/Low | 165/1172 | 1.00 (reference) |  | 613/1670 | 1.62 (1.38, 1.91) |
|  | Moderate | 71/515 | 0.97 (0.74, 1.26) |  | 155/389 | 1.64 (1.35, 2.00) |
|  | High | 38/143 | 1.53 (1.12, 2.10) |  | 76/145 | 1.99 (1.58, 2.50) |
| RRs (95% CI) for moderate-risk alcohol consumption within strata of physical activity |  |  | 0.97 (0.74, 1.26) |  |  | 1.01 (0.87, 1.17) |
| RRs (95% CI) for high-risk alcohol consumption within strata of physical activity |  |  | 1.53 (1.12, 2.10) |  |  | 1.23 (1.02, 1.47) |
| Measure of interaction on additive scale (moderate): 0.05 (-0.30, 0.40), p=0.7706 | | | | | | |
| Measure of interaction on additive scale (high): -0.17 (-0.75, 0.42), p=0.5769 | | | | | | |
| Measure of interaction on multiplicative scale (moderate): 1.05 (0.77, 1.41), p=0.7700 | | | | | | |
| Measure of interaction on multiplicative scale (high): 0.80 (0.56, 1.15), p=0.2298 | | | | | | |

*RRs adjusted for age, gender, race/ethnicity, education, body mass index, relationship status, or any chronic conditions*

| **Supplementary Table S5. Effect modification of the association between alcohol consumption risk and anxiety/depression by physical activity level (≥150 vs. <150 mins/week) among participants without chronic conditions or poor health, CHASING COVID Cohort, September 2021–December 2023 (N=3,141)** | | | | | | |
| --- | --- | --- | --- | --- | --- | --- |
|  |  | Meets physical activity recommendations  (≥ 150 mins/wk) | |  | Does not meet physical activity recommendations (< 150 mins/wk) | |
|  |  | *N* with/without anxiety/depression | RR (95% CI); *P* |  | *N* with/without anxiety/depression | RR (95% CI); *P* |
| Alcohol consumption risk level | None/Low | 116/798 | 1.00 (reference) |  | 278/942 | 1.38 (1.12, 1.69) |
|  | Moderate | 54/372 | 0.97 (0.71, 1.32) |  | 90/248 | 1.62 (1.26, 2.07) |
|  | High | 20/101 | 1.16 (0.74, 1.82) |  | 36/86 | 1.81 (1.30, 2.52) |
| RRs (95% CI) for moderate-risk alcohol consumption within strata of physical activity |  |  | 0.97 (0.71, 1.32) |  |  | 1.17 (0.96, 1.44) |
| RRs (95% CI) for high-risk alcohol consumption within strata of physical activity |  |  | 1.16 (0.74, 1.82) |  |  | 1.31 (0.98, 1.77) |
| Measure of interaction on additive scale (moderate): 0.27 (-0.15, 0.69), p=0.2089 | | | | | | |
| Measure of interaction on additive scale (high): 0.28 (-0.45, 1.00), p=0.4554 | | | | | | |
| Measure of interaction on multiplicative scale (moderate): 1.21 (0.84, 1.74), p=0.3071 | | | | | | |
| Measure of interaction on multiplicative scale (high): 1.14 (0.66, 1.95), p=0.6444 | | | | | | |

*RRs adjusted for age, gender, race/ethnicity, education, body mass index, relationship status*

| **Supplementary Table S6. Cohort characteristics by modifier status (sensitivity definition), CHASING COVID Cohort, September 2021-December 2023 (N=5,152)** | | | | |
| --- | --- | --- | --- | --- |
|  | Total | Physical activity | | |
|  |  | ≥ 150 mins/wk | < 150 mins/wk |  |
|  | N (%) | n (col %) | n (col %) | ꭕ^2^ p-value |
| Total | 5152 (100) | 2104 (40.8) | 3048 (59.2) |  |
| Alcohol consumption level |  |  |  | <0.0001 |
| None/low-risk | - | 1337 (63.6) | 2283 (74.9) |  |
| Moderate-risk | - | 586 (27.9) | 544 (17.9) |  |
| High-risk | - | 181 (8.6) | 221 (7.3) |  |
| Age |  |  |  | <0.0001 |
| 18-29 | 1129 (21.9) | 402 (19.1) | 727 (23.9) |  |
| 30-39 | 1486 (28.8) | 568 (27.0) | 918 (30.1) |  |
| 40-49 | 945 (18.3) | 364 (17.3) | 581 (19.1) |  |
| 50-64 | 699 (13.6) | 294 (14.0) | 405 (13.3) |  |
| 65+ | 893 (17.3) | 476 (22.6) | 417 (13.7) |  |
| Sex |  |  |  | <0.0001 |
| Male | 2337 (45.4) | 1025 (48.7) | 1790 (58.7) |  |
| Female | 2815 (54.6) | 1079 (51.3) | 1258 (41.3) |  |
| Race/ethnicity |  |  |  | <0.0001 |
| Hispanic | 849 (16.5) | 301 (14.3) | 548 (18.0) |  |
| White, non-Hispanic | 3225 (62.6) | 1426 (67.8) | 1799 (59.0) |  |
| Black, non-Hispanic | 547 (10.6) | 153 (7.3) | 394 (12.9) |  |
| Asian/Pacific Islander | 368 (7.1) | 164 (7.8) | 204 (6.7) |  |
| Other, non-Hispanic | 163 (3.2) | 60 (2.9) | 103 (3.4) |  |
| Education |  |  |  | <0.0001 |
| High school or less | 609 (11.8) | 111 (5.3) | 498 (16.3) |  |
| Some college | 1350 (26.2) | 390 (18.5) | 960 (31.5) |  |
| College graduate | 3193 (62.0) | 1603 (76.2) | 1590 (52.2) |  |
| Annual household income |  |  |  | <0.0001 |
| <$35,000 | 1426 (27.7) | 359 (17.1) | 1067 (35.0) |  |
| $35,000-$49,999 | 600 (11.7) | 191 (9.1) | 409 (13.4) |  |
| $50,000-$69,999 | 771 (15.0) | 326 (15.5) | 445 (14.6) |  |
| $70,000-$99,999 | 879 (17.1) | 402 (19.1) | 477 (15.7) |  |
| $100,000 | 1476 (28.7) | 826 (39.3) | 650 (21.3) |  |
| BMI category |  |  |  | <0.0001 |
| Underweight | 107 (2.1) | 40 (2.0) | 67 (2.3) |  |
| Normal | 1873 (37.4) | 927 (45.4) | 946 (32.0) |  |
| Overweight | 1505 (30.1) | 666 (32.6) | 839 (28.3) |  |
| Obesity | 1519 (30.4) | 411 (20.1) | 1108 (37.4) |  |
| In a relationship^§§^ |  |  |  | <0.0001 |
| Yes | 3192 (62.7) | 1399 (66.8) | 1793 (59.8) |  |
| No | 1901 (37.3) | 695 (33.2) | 1206 (40.2) |  |
| Chronic health condition dx or poor general health |  |  |  | <0.0001 |
| Yes | 2011 (38.6) | 643 (30.6) | 1368 (44.8) |  |
| No | 3141 (61.4) | 1461 (69.4) | 1680 (55.1) |  |
| Ever in recovery from drugs and/or alcohol |  |  |  | <0.0001 |
| Yes | 729 (14.2) | 532 (17.5) | 197 (9.4) |  |
| No | 4422 (85.8) | 2515 (82.5) | 1907 (90.6) |  |

‡Chi-squared test used for assessment of statistically significant differences across the three groups
^§^<1% missing ^§§^ 1-3% missing; ^§§§^ 4-10% missing

| **Supplementary Table S7. Anxiety and/or depression (outcome) by alcohol consumption level (exposure) and physical activity level (modifier, sensitivity definition), CHASING COVID Cohort, September 2021-December 2023** | | | | | | | | |
| --- | --- | --- | --- | --- | --- | --- | --- | --- |
| Outcome Variable | Total N, % | Alcohol consumption level, outcome (N, column %) | | | | Physical activity, modifier (N, column %) | | |
|  |  | None/low | Moderate | High | *ꭕ^2^ p-val* | ≥ 150 mins/wk | < 150 mins/wk | *ꭕ^2^ p-val* |
| Full Cohort (N=5,152) |  |  |  |  |  |  |  |  |
| Total | 5152 (100) | 3620 (70.3) | 1130 (21.9) | 402 (7.8) |  | 2104 (40.8) | 3620 (70.3) |  |
| Anxiety and/or depression |  |  |  |  | 0.0019 |  |  | <0.0001 |
| Yes | 1118 (21.7) | 778 (21.5) | 226 (20.0) | 114 (28.4) |  | 274 (13.0) | 778 (21.5) |  |
| No | 4034 (78.3) | 2842 (78.5) | 904 (80.0) | 288 (71.6) |  | 1830 (87.0) | 2842 (78.5) |  |
| Subsample Without Underlying Condition(s) (N=3,141) |  |  |  |  |  |  |  |  |
| Total | 3141 (100) | 2134 (67.9) | 764 (24.3) | 243 (7.7) |  | 1461 (56.5) | 1680 (53.5) |  |
| Anxiety and/or depression |  |  |  |  | 0.2243 |  |  | <0.0001 |
| Yes | 594 (18.9) | 394 (18.5) | 144 (18.9) | 56 (23.1) |  | 190 (13.0) | 404 (24.1) |  |
| No | 2547 (81.1) | 1740 (81.5) | 620 (81.2) | 187 (77.0) |  | 1271 (87.0) | 1276 (76.0) |  |

| **Supplementary Table S8. Comparison of cohort characteristics for the full cohort and the analytic sample, CHASING COVID Cohort** | | |
| --- | --- | --- |
|  | Full sample (N=5,798) | Analytic Sample (N=5,152) |
|  | n (%) | n (%) |
| Age |  |  |
| 18-29 | 1326 (22.9) | 1129 (21.9) |
| 30-39 | 1680 (29.0) | 1486 (28.8) |
| 40-49 | 1051 (18.1) | 945 (18.3) |
| 50-64 | 754 (13.0) | 699 (13.6) |
| 65+ | 987 (17.0) | 893 (17.3) |
| Sex |  |  |
| Male | 2652 (46.0) | 2337 (45.4) |
| Female | 3116 (54.0) | 2815 (54.6) |
| Race/ethnicity |  |  |
| Hispanic | 1001 (17.3) | 849 (16.5) |
| White, non-Hispanic | 3545 (61.1) | 3225 (62.6) |
| Black, non-Hispanic | 662 (11.4) | 547 (10.6) |
| Asian/Pacific Islander | 404 (7.0) | 368 (7.1) |
| Other, non-Hispanic | 186 (3.2) | 163 (3.2) |
| Education |  |  |
| High school or less | 733 (12.6) | 609 (11.8) |
| Some college | 1576 (27.2) | 1350 (26.2) |
| College graduate | 3489 (60.2) | 3193 (62.0) |
| Annual household income |  |  |
| <$35,000 | 1657 (28.6) | 1426 (27.7) |
| $35,000-$49,999 | 669 (11.5) | 600 (11.7) |
| $50,000-$69,999 | 857 (14.8) | 771 (15.0) |
| $70,000-$99,999 | 958 (16.5) | 879 (17.1) |
| $100,000 | 1653 (28.5) | 1476 (28.7) |
| BMI category |  |  |
| Underweight | 127 (2.3) | 107 (2.1) |
| Normal | 2103 (37.6) | 1873 (37.4) |
| Overweight | 1684 (30.1) | 1505 (30.1) |
| Obesity | 1686 (30.1) | 1519 (30.4) |
| In a relationship^§§^ |  |  |
| Yes | 3231 (62.6) | 3192 (62.7) |
| No | 1927 (37.4) | 1901 (37.3) |
| Chronic health condition dx or poor general health |  |  |
| Yes | 2264 (39.1) | 2011 (38.6) |
| No | 3534 (61.0) | 3141 (61.4) |
| Ever in recovery from drugs and/or alcohol |  |  |
| Yes | 733 (14.2) | 729 (14.2) |
| No | 4435 (85.8) | 4422 (85.8) |
